# The diagnostic role of abductor pollicis brevis -first dorsal interosseous (APB-FDI) index in diagnosing Amyotrophic lateral sclerosis (ALS) and comparison of its diagnostic performance with other tests demonstrating split hand phenomenon among Bangladeshi population

**DOI:** 10.1101/2023.08.04.23293649

**Authors:** Md. Shuktarul Islam, Hashmi Sina, Iftikher Alam, Md. Arifuzzaman, Reaz Mahmud

## Abstract

**Background:** The split hand phenomenon is an early and distinctive feature of amyotrophic lateral sclerosis (ALS), resulting from the differential wasting of the intrinsic hand muscles, particularly the thenar muscles. This study aimed to assess the diagnostic role of the APB-FDI Index in the diagnosis of Amyotrophic Lateral Sclerosis (ALS) among the Bangladeshi population. We also compared its diagnostic performances with FDI-ADM and ADM-APB ratios measuring the split hand phenomenon.

**Methods:** We carried out this cross-sectional study in the Department of Neurology, Sir Salimullah Medical College Mitford Hospital, and Dhaka Medical College Hospital, Dhaka, from March 2019 to September 2021. We determined the APB-FDI index with APB (CMAP) X FDI (CMAP) /ADM (CMAP). We also compared the findings with age and gender-matched healthy control and disease control (Hirayama disease). We studied the value of using APB-FDI, FDI-ADM, and ADM-APB ratios for diagnosis. We analyzed the data with ROC curves and calculated AUC for each ratio. We also used IDI and DCA to compare diagnostic accuracy.

**Results:** We studied 43 people with ALS, 30 healthy people, and 10 Hirayama patients. Most ALS patients had cervical phenotypes (31, 72%) and a median ALS functional rating score of 27(IQR,25-32). The cutoff values of APB-FDI Index, ADM-APB CMAP amplitude ratio, and FDI-ADM CMAP amplitude ratio are 4.27, 2.20 respectively. The APB-FDI Index has a high AUC of 0.9, with good sensitivity and specificity. However, for Hirayama disease, the APB-FDI Index cutoff value is 11.6. The APB-FDI Index is clinically effective with optimal threshold probabilities identified through the Youden-Index ((APB-FDI Index-0.72, ADM-APB CMAP amplitude ratio-0.51, and FDI-ADM CMAP amplitude ratio-0.17). Its standardized net benefits exceed healthy control and Hirayama when risk thresholds are between 0.1 and 0.8. The IDI results showed that the APB-FDI Index was superior to the ADM-APB CMAP amplitude ratio and FDI-ADM CMAP amplitude ratio in healthy control and Hirayama although p value is insignificant.

**Conclusion:** APB-FDI Index is the sensitive, specific, and early diagnostic marker for ALS. Its diagnostic performance is better than the other tests.

## Introduction

Amyotrophic lateral sclerosis (ALS) is a progressive degenerative disorder [1] involving the motor neurons, a heterogeneous disease that begins focally and becomes widespread within a short period. The patient dies within three years of symptom onset due to respiratory muscle involvement [2]. The mechanisms of neuronal degeneration include oxidative stress, excitotoxicity, mitochondrial dysfunction, glial activation, RNA processing, and growth factor abnormalities. They may occur parallelly or in sequence [3]. There is no definitive diagnostic tool for diagnosing ALS. The progressive upper motor neuron (UNM) and lower motor neuron (LMN) findings in the history and examination may diagnose ALS with 95% accuracy [4]. For the uniformity of the diagnosis, El Escorial criteria were developed [5,6]. The sensitivity of these criteria is low in case of early stages of the disease [7]. Possible differentials like Spinal muscular atrophy (SMA), Hirayama disease, cervical myeloradiculopathy, and Multifocal motor neuropathy with conduction block (MMCB) are sometimes difficult to exclude [8]. The median diagnostic time from the symptom onset is 11.5 months, and about half of the patients received alternative diagnoses [9]. A dissociated pattern of muscle atrophy, first described as a split hand by Wilbourn [10], was observed in 60-70 % of ALS patients [9,11]. Here the abductor pollicis brevis (APB) and first dorsal interosseous (FDI) muscles are involved more than the hypothenar muscles. The cause of differential muscle involvement in ALS is unknown; there are theories for cortical and peripheral mechanisms.[12] Among the postulated peripheral mechanism, higher nodal persistence of sodium currents in the APB and FDI than in ADM might be a mechanism [13]. Furthermore, the cortical mechanism involves preferential involvement of corticomotoneuronal input to the thenar muscles [14]. In our day-to-day activities, using a pincer grasp is essential (adductor pollicis, first dorsal interosseous and flexor pollicis brevis muscles.), making the spinal motor neurons innervating these muscles liable to more oxidative stress.

The corticospinal connections to FDI far outnumber those of ADM. It may produce more glutamate excitotoxicity in the FDI spinal neurons [15]. The APB-FDI index quantifies this preferential muscle involvement, which is derived by multiplying the APB and FDI CMAP amplitudes and dividing the product by the ADM CMAP amplitude according to the following formula: 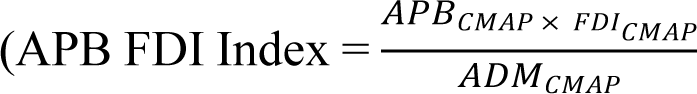 [16]. This index helps detect the early stages of ALS, not fulfilling the diagnostic criteria [11]. This phenomenon did not differ in different stages and certainty [17]. The cutoff value of this index is 5.2 and exhibits 74% sensitivity and 80% specificity in diagnosing ALS [11]. It is more sensitive than the two other ratios used to quantify the split hand phenomenon (ADM/APB CMAP amplitude ratio >1.7, 51% sensitive, and FDI/ADM less than 0.9 is 34 % sensitive) [16]). This method was not examined among the Bangladeshi population, and the cutoff value is largely unknown among the Bangladeshi people. We planned to assess the diagnostic role of the APB FDI Index, ADM/APB CMAP amplitude ratio, and FDI/ADM CMAP amplitude ratio in the diagnosis of Amyotrophic Lateral Sclerosis (ALS) and to determine their cutoff values for the Bangladeshi population. We also compared their diagnostic performance in diagnosing ALS.

## Methods

This cross-sectional study was done in the Department of Neurology at Sir Salimullah Medical College Mitford Hospital, Dhaka, and Dhaka Medical College from March 2019 to September 2020. We obtained Ethical clearance from the ethical review committee of Sir Salimullah Medical College Mitford Hospital (SSMC/2019/224).

### Participants

We enrolled all the probable, possible, and definite cases of Amyotrophic lateral sclerosis (ALS), according to the revised El Escorial Diagnostic Criteria for ALS [5], presented in electrophysiology lab of the neurology department of Sir Salimullah Medical College Mitford Hospital and Dhaka Medical College. Patients aged 18 years or more and of both sexes were included in this study. Patients with undetectable CMAP amplitude of any muscle (APB, FDI, and ADM), associated radiculopathy, peripheral neuropathy, or neuromuscular disease were excluded from the study. We recruited healthy volunteers as the control. We also took patients with Hirayama disease, diagnosed according to criteria outlined by Tashiro et al[18]. and confirmed with MRI cervical spine flexion and extension view as disease control. Both the cases and control participants gave Informed written consent. The calculated sample size was 30 for ALS and healthy-control, considering the abnormal split hand index proportion of 41% in the case and 5% in the healthy control group, according to Kuwabara et al. 2008[19]. With a 5% level of significance in the two-tailed test and 90% power. We used the following formula to calculate the sample size 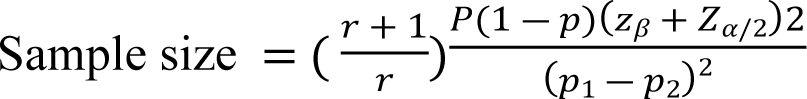 (Z_α_ =1.96 at 5% level of significance, Z_β_=1.28 at 90% power, P_1_=0.41 P_2_=0.05, P= (p_1_+p_2_) ÷2=0.23).

We also compared the electrophysiology findings of 10 Hirayama patients with the ALS patients.

### Study procedure

The demographic profile and complaints were recorded in a preformed case record form. The expert neurologist did detail Neurologic examinations. The hemoglobin, total count, differential count, blood sugar, serum creatinine, brain CT scan, and MRI cervical spine were done to exclude the other diagnoses for all the included patients. Revised El Escorial Diagnostic Criteria for ALS diagnose patients with ALS [5]. The disability of ALS was assessed with ALS Functional Rating Scale-Revised (ALS-FRS-R) [20]. The Hirayama disease was diagnosed with Tashiro criteria [18].

### NCS assessment

The NCS was done with NIHON KOHDEN Neuropack MEB-7102 mobile unit with a two-channel evoked potential/EMG measuring system (Nihon Kohden Corporation, Tokyo, Japan) according to the American Association of Electrodiagnostic Medicine, American Academy of Neurology, and American Academy of Physical Medicine and Rehabilitation guidelines. We maintained the temperature at 33-34 degrees centigrade. We did the neurophysiological study on the most affected hand and opposite lower limb in cervical onset MND, including the measurement of motor conduction velocity (MCV), distal motor latency (DML), and compound muscle action potential amplitude (CMAP), F-latency, sensory conduction velocity (SCV), distal latency and sensory nerve action potential amplitude (SNAP) of the examined nerves. In limb onset type MND, we did the electrophysiological study on the most affected lower limb and opposite upper limb, and in bulbar onset MND in the right hand and left leg.

Motor nerve conduction studies were performed on the median and ulnar nerves with compound muscle action potential (CMAP) responses recorded from the abductor pollicis brevis (APB), first dorsal interosseous (FDI), and abductor digit minimi (ADM) muscles. The Responses were recorded by 10 mm gold disc electrodes positioned in a belly tendon arrangement over each muscle. We Placed the active electrode over the midpoint of the respective muscle, ensuring a negative take-off of the CMAP response, while the reference electrode was positioned over the base of the thumb (APB and FDI CMAP recordings) and base of digit 5 (for ADM CMAP recordings). We Set the distance between the stimulating cathode and active electrode for APB and ADM, CMAP to 5 cm, and for FDI CMAP to 8 cm. We set the filter at 3 and 10 kHz, the sweep speed at 20 msec, or 2 msec/division, and the sensitivity for recording CMAP responses at 5 mV. The pulse duration was 0.2 ms. We increased an additional 25% in the current to ensure the Supramaximal stimulation. Measurements of the compound muscle action potential (CMAP) included latency (onset and peak) and conduction velocity (CV). In the study, we used belly tendon montage, gave the stimulation at the wrist using an orthodromic technique with a bipolar stimulator, and expressed the distal latency as a millisecond and conduction velocity as a meter per second.

We Calculated the APB FDI index by multiplying the CMAP amplitude recorded over the APB and FDI muscles and dividing this product by the CMAP amplitude recorded over the ADM muscle, as follows: APB FDI sindex = APB CMAP X FDI CMAP /ADM CMAP. We also calculated ADM/APB CMAP amplitude ratio, and FDI/ADM CMAP amplitude ratio. For the analysis, we took the values from the most affected hand in the case of ALS and Hirayama disease. In the case of control, we generally use right-hand parameters.

### Statistical analysis

Data were analyzed using SPSS 20 and R (v4.1.1). Qualitative data were expressed in number, percentage, normally distributed quantitative data as mean (SD), and non-normal data as median (IQR). Comparisons between groups (continuous parameters) were made by Student’s t-test and skewed parameters with Mann Whitney U test. The Chi-Square test compared categorical parameters. Several indicators were used to measure the performance of the APB-FDI Index, FDI-ADM, and ADM-APB ratios. The prediction scoring systems were analyzed using the receiver operating characteristic (ROC) curve and the area under the ROC curves (AUC). DeLong’s test was used to determine the statistical difference between AUCs. The Integrated Discrimination Improvement (IDI) indicator was also used to evaluate their performance. Decision Curve Analysis (DCA) was also performed to assess the clinical usefulness of these ratios. The significance of the results, as determined by a 95% confidence interval and a value of *p*<0.05, was considered statistically significant.

## Results

This cross-sectional study was conducted in the Neurology department of Sir Salimullah Medical College Mitford Hospital and Dhaka Medical College, Dhaka, from March 2019 to March 2022. We screened 57 suspected patients with ALS and included 43 patients after considering the patient’s inclusion, exclusion criteria, and consent. We had 30 age and sex-matched healthy control (Figure 1). We also included 10 Hirayama patients to compare the index and the ratios (APB-FDI Index, ADM-APB CMAP amplitude ratio, and FDI-ADM CMAP amplitude ratio)

**Figure 1:**
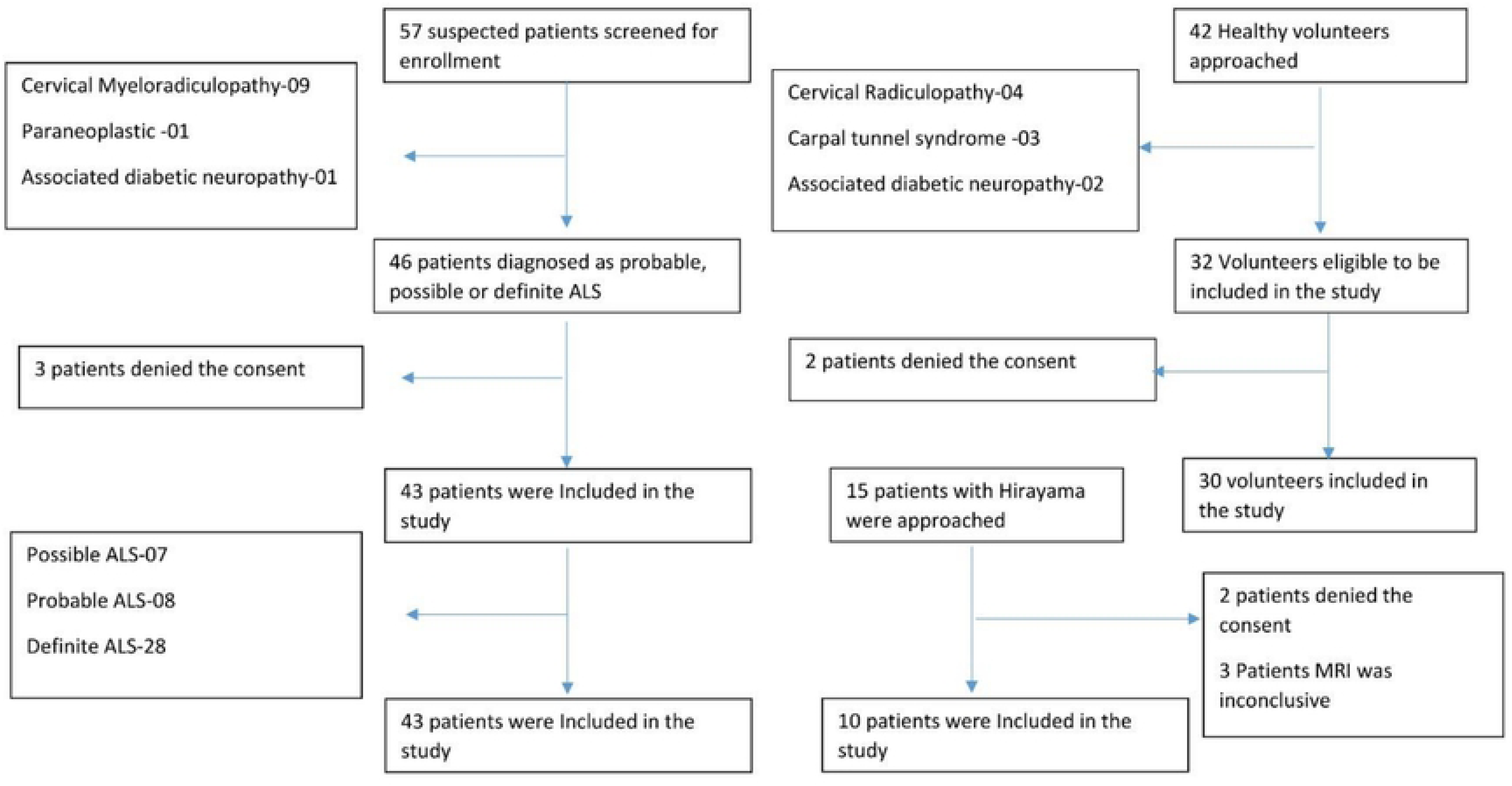
Patients and volunteer selection for this study. We screened 57 suspected patients with Motor Neuron Disease and included 43 patients after screening. We had 30 healthy volunteers and ten patients with Hirayama disease for comparison

### Demographic and clinical profile of the participants

The age of the ALS group was generally >40 years (88.4%), and the mean age (SD) was 56.1(14.8). The age of patients with ALS was comparable with the control. Whereas the Hirayama patients were generally < 40 years (90%), and the mean age was 24.7(13.7). The males were similarly frequent in ALS, 34(79.1), and Hirayama groups, 7(70%). 15(34.4%) of the ALS patients were underweight. The mean (SD) weight of the ALS and Hirayama was 18.8(1.8) and 21.6(2.2), respectively (Table 1).

**Table 1:** Demographic characteristics of the study participants

| Trait | Group I<br>(cases)<br>N=43 | Group II(Control)<br>N=30 | Group III<br>(Hirayama)<br>N=10 |
| --- | --- | --- | --- |
| Age in years, mean (SD) | 56.1(14.8) | 57.7(9.7) | 24.7(13.7) |
| Age groups, n (%) |  |  |  |
| <40 years | 5(11.6) | 2(6.7) | 9(90) |
| 40-60 years | 22(51.2) | 21(70) | 0(0) |
| >60 years | 16(37.2) | 7(23.3) | 1(10) |
| Sex (Male), n (%) | 34(79.1) | 22(73.3) | 7(70) |
| BMI, Mean (SD) | 18.8(1.8) | 22.8(1.65) | 21.6(2.2) |
| BMI groups, n (%) |  |  |  |
| Underweight (<18.5 kg/m<sup>2</sup>) | 15(34.9) | 0(0) | 1(10) |
| Normal weight (18.5-22.9 kg/m<sup>2</sup>) | 21(48.8) | 18(60) | 7(70) |
| Overweight (23-24.9 kg/m<sup>2</sup>) | 6(14) | 11(36.7) | 1(10) |
| Obesity (25-29.9 Kg/m<sup>2</sup>) | 1(2.3) | 1(3.3) | 1(10) |

In this study total of 28(65.1) had definite ALS and presented with median (IQR), 6(5-10) months of illness. They were generally cervical phenotype, 31(72.1%), and they had ALS functional rating score, median (IQR) of 27(25-32). (Table 2).

**Table 2:** Clinical characteristics of the ALS patients

| Clinical Characteristics | Results |
| --- | --- |
| **Disease Categories according to El scorial criteria, n (%)** |  |
| Possible | 7(16.3) |
| Probable | 8(18.6) |
| Definite | 28(65.1) |
| Age of onset, Median (IQR) | 57(46-64) |
| Phenotype according to the onset |  |
| Bulbar | 3(7) |
| Cervical | 31(72.1) |
| Lumbar | 2(4.7) |
| Mixed | 7(16.3) |
| Disease duration, months, median (IQR) | 6(5-10) |
| ALS function rating score, median (IQR) | 27(25-32) |

The median (IQR) CMAP amplitude of FDI, APB, and ADM, 2.8(0.8-5.3), 2.3(0.8-4.6), and 3.8(1.8-6.9) respectively were significantly low (p-value <0.001) than the healthy control. Among ALS patients amplitude of FDI and APB is lower than the amplitude of ADM. The median (IQR) of APB-FDI index, APB-ADM ratio and FDI-ADM ratio were 1.7(0.3-3.8), 0.6(0.3-1.1) and 0.8(0.4-1.1) were significantly lower than that of healthy control (p value<0.001, 0.016, 0.003 respectively). The ADM-APB ratio of 1.7(0.9-3.2) is significantly higher than the healthy control (p-value 0.016). (Table 3)

**Table 3:**
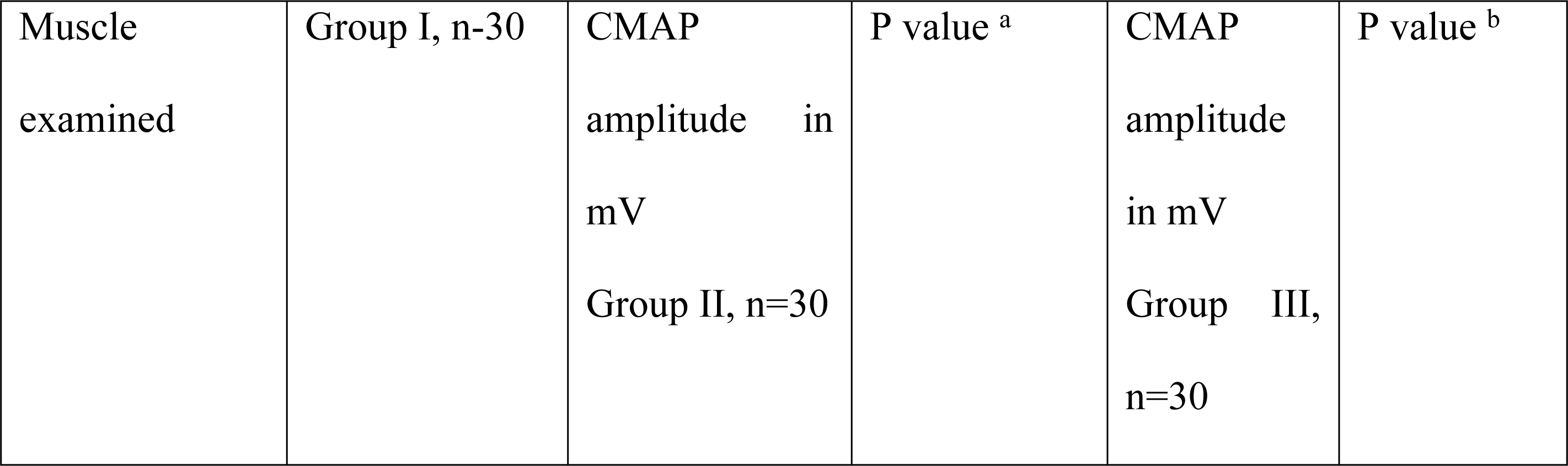
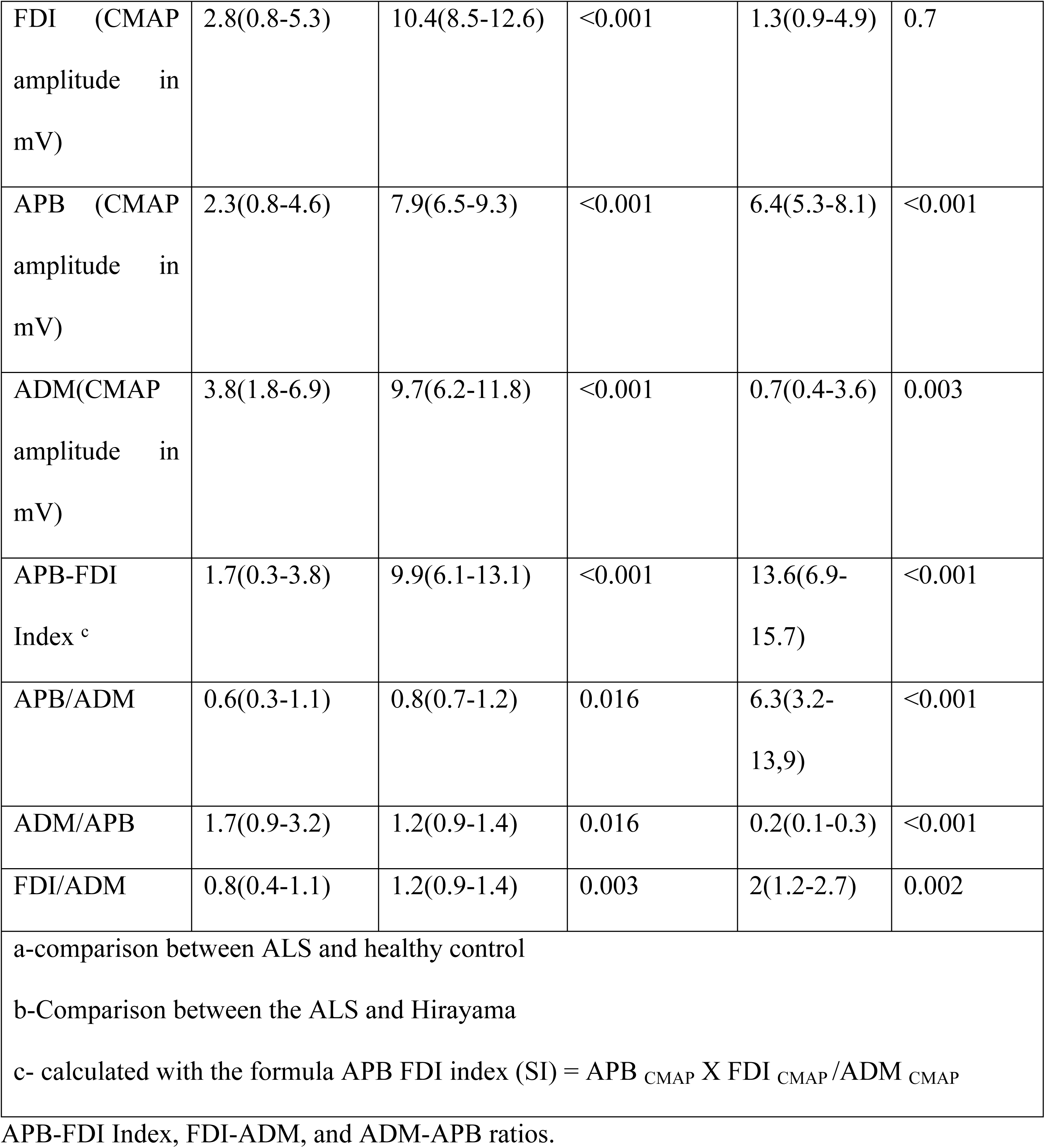
Comparison of Electrophysiologic findings with the healthy controls and Hirayama disease

| Muscle<br>examined | Group I, n=30 | CMAP<br>amplitude in<br>mV<br>Group II, n=30 | P value <sup>a</sup> | CMAP<br>amplitude<br>in mV<br>Group III,<br>n=30 | P value <sup>b</sup> |
| --- | --- | --- | --- | --- | --- |
| FDI (CMAP amplitude in mV) | 2.8(0.8-5.3) | 10.4(8.5-12.6) | <0.001 | 1.3(0.9-4.9) | 0.7 |
| APB (CMAP amplitude in mV) | 2.3(0.8-4.6) | 7.9(6.5-9.3) | <0.001 | 6.4(5.3-8.1) | <0.001 |
| ADM(CMAP amplitude in mV) | 3.8(1.8-6.9) | 9.7(6.2-11.8) | <0.001 | 0.7(0.4-3.6) | 0.003 |
| APB-FDI Index <sup>c</sup> | 1.7(0.3-3.8) | 9.9(6.1-13.1) | <0.001 | 13.6(6.9-15.7) | <0.001 |
| APB/ADM | 0.6(0.3-1.1) | 0.8(0.7-1.2) | 0.016 | 6.3(3.2-13,9) | <0.001 |
| ADM/APB | 1.7(0.9-3.2) | 1.2(0.9-1.4) | 0.016 | 0.2(0.1-0.3) | <0.001 |
| FDI/ADM | 0.8(0.4-1.1) | 1.2(0.9-1.4) | 0.003 | 2(1.2-2.7) | 0.002 |
| a-comparison between ALS and healthy control |  |  |  |  |  |
| b-Comparison between the ALS and Hirayama |  |  |  |  |  |
| c- calculated with the formula APB FDI index (SI) = APB<sub>CMAP</sub> X FDI<sub>CMAP</sub>/ADM<sub>CMAP</sub> |  |  |  |  |  |

APB-FDI Index, FDI-ADM, and ADM-APB ratios.

CMAP amplitude of the FDI did not differ between the ALS and Hirayama patients (p-value 0.7). Whereas the median (IQR) amplitude of the APB, 6.4(5.3-8.1), is significantly higher (p-value <0.001), and the amplitude of ADM, 0.7(0.4-3.6) was significantly lower (p value0.003) in the Hirayama than ALS.

The median (IQR) of APB-FDI index, and FDI-ADM ratio was 13.6(6.9-15.7), and 2(1.2-2.7) were significantly higher in Hirayama group than that of healthy control (p value<0.001, <0.001, 0.002 respectively). Whereas the ADM-APB ratio of 0.2(0.1-0.3) is significantly lower in the Hirayama group than in the ALS group (p-value <0.001).

There was no correlation between the disease severity of ALS and the APB-FDI index (r= -0.16, p-value 0.28) (Supplement 1).

### Comparison of ROC curves

To diagnose ALS, the APB-FDI Index, ADM-APB CMAP amplitude ratio, and FDI-ADM CMAP amplitude ratio are used with cutoff values of 4.27, 1.04, and 2.20, respectively. The APB-FDI Index has a high sensitivity and specificity with an AUC of 0.9. However, when differentiating Hirayama disease, the APB-FDI Index’s cutoff value is higher at 11.6, but its specificity is lower at 0.7. In comparison, the APB-FDI Index is the most effective in diagnosing ALS compared to healthy individuals, but it has lower performance in differentiating Hirayama disease (Figure 2).

**Figure 2:**
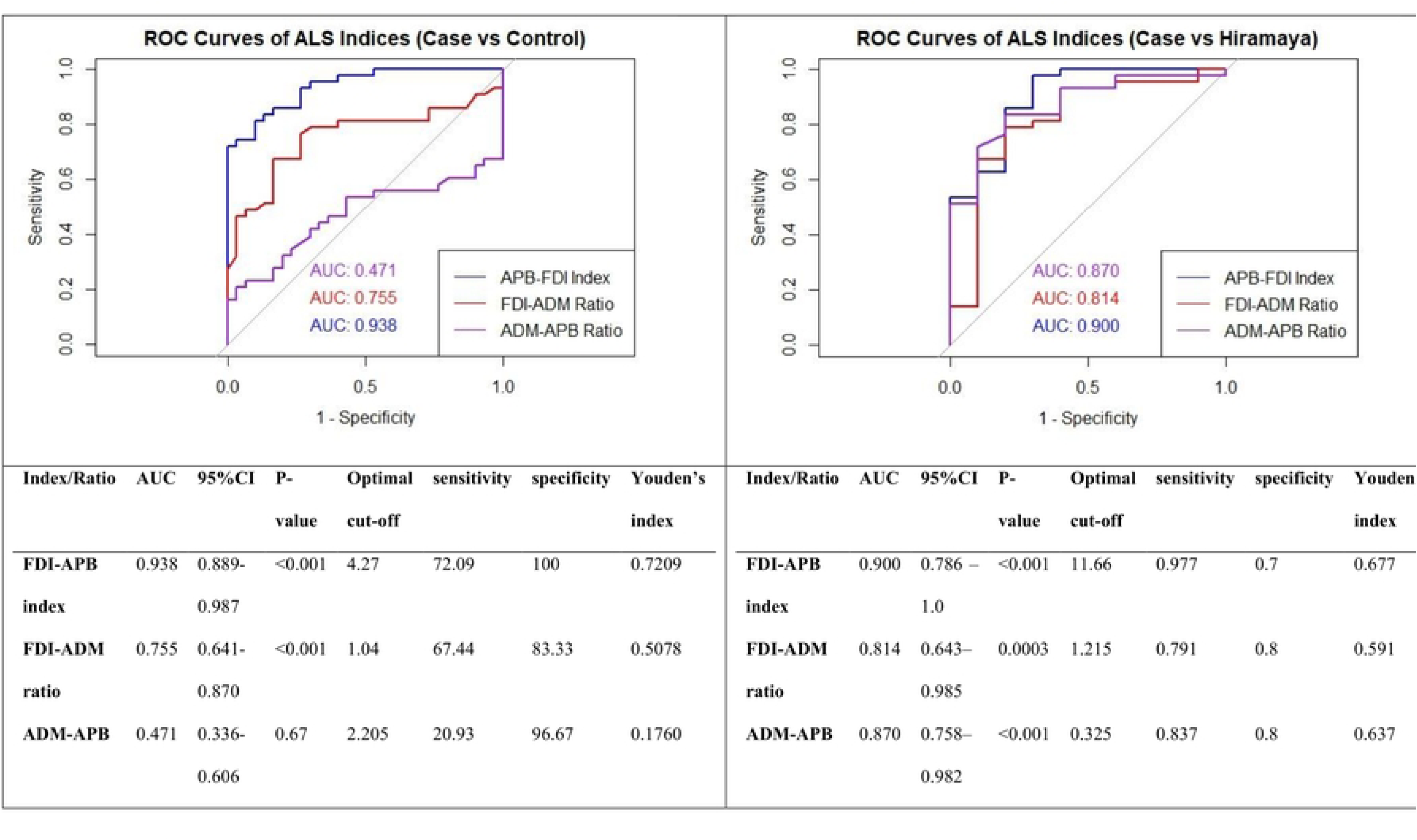
Comparison of the ROC curve and diagnostic performances of APB-FD**I** Index, ADM-APB CMAP amplitude ratio, and FDI-ADM CMAP amplitude ratio

### Comparison of IDI

We evaluated the prediction performance of the scoring systems by calculating IDIs, corresponding P values, and confidence intervals using 1000 times bootstrap resampling (Table 4). The IDI results showed that the APB-FDI Index was superior to the ADM-APB CMAP amplitude ratio and FDI-ADM CMAP amplitude ratio in healthy control and Hirayama although p value is insignificant. The IDI for APB-FDI Index in comparison to ADM-APB CMAP amplitude ratio and FDI-ADM CMAP amplitude ratio was 0.55 (95% CI -0.661, -0.434; p=0.481) and -0.55 (95% CI -0.658, -0.440; p=0.504), respectively in healthy control. In Hirayama, the IDI for APB-FDI Index in comparison to ADM-APB CMAP amplitude ratio and FDI-ADM CMAP amplitude ratio was -0.228 (95% CI -0.416, -0.015; p=0.487) and -0.368 (95% CI -0.541, -0.172; p=0.465), respectively.

**Table 4.** Pairwise comparison of IDIs in overall patients

| **Comparison of models**<br>**(New model VS reference model)** | **IDI** | **95% CI of IDI** | **P-value of IDI** |
| --- | --- | --- | --- |
| **For healthy control** |  |  |  |
| **FDI-APB index vs FDI-ADM ratio** | **-0.549** | **-0.658, -0.440** | **0.504** |
| **FDI-APB index vs ADM-APB** | **-0.545** | **-0.661, -0.434** | **0.481** |
| **FDI-ADM ratio vs ADM-APB** | **0.007** | **-0.035, 0.051** | **0.495** |
| **For Hirayama** |  |  |  |
| **FDI-APB index vs FDI-ADM ratio** | **-0.368** | **-0.541, -0.172** | **0.465** |
| **FDI-APB index vs**<br>**ADM-APB** | **-0.228** | **-0.416, -0.015** | **0.487** |
| **FDI-ADM ratio vs**<br>**ADM-APB** | **0.133** | **-0.028, 0.262** | **0.495** |

### Comparison of DCA curves

The results of DCA (Fig 3) demonstrate the clinical value of evaluating standardized net benefits (sNBs) and risk threshold probabilities using the APB-FDI Index, ADM-APB CMAP amplitude ratio, and FDI-ADM CMAP amplitude ratio. The sNBs of the APB-FDI Index were higher than the extreme curve for healthy control and Hirayama when the range of risk threshold probabilities was between 0.1 and 0.8. Compared to the ADM-APB CMAP and FDI-ADM CMAP amplitude ratios, the APB-FDI Index had higher sNBs when the range of threshold probabilities was between 0.1 and 0.8, indicating its broad clinical utility. Optimal threshold probabilities were also determined for each index using the Youden-Index (APB-FDI Index-0.72, ADM-APB CMAP amplitude ratio-0.51, and FDI-ADM CMAP amplitude ratio-0.17), and the DCA results show that the APB-FDI Index had higher sNBs than the other two ratios. Furthermore, the results and conclusions remained consistent for threshold probabilities between 0.45 and 0.80.

**Figure 3.**
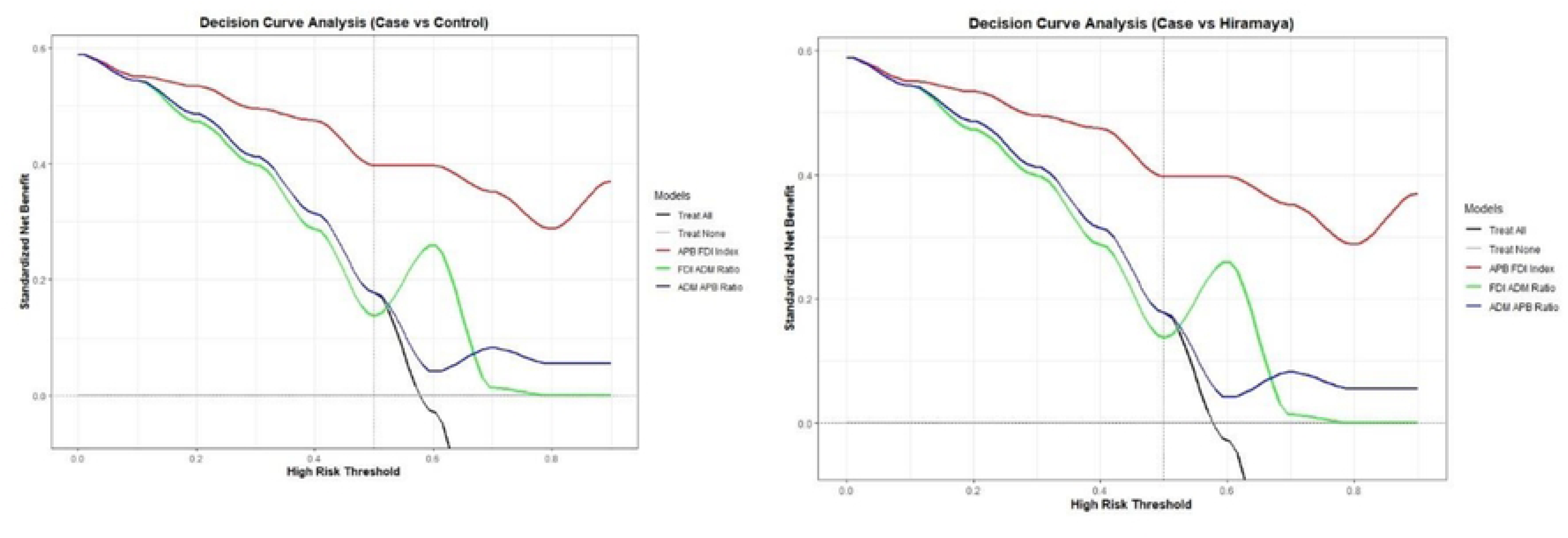
DCA curves of APB-FDI Index, ADM-APB CMAP amplitude ratio, and FDI-ADM CMAP amplitude ratio

## Discussion

Our study confirms the split hand phenomenon in ALS patients, where the CMAP amplitude of the ADM muscle is higher than that of the FDI and APB muscles. We measured this phenomenon using the APB-FDI index, APB-ADM ratio, and FDI-ADM ratio. The cutoff values for these measurements are 4.27, 1.04, and 2.20, respectively. The APB-FDI index is more sensitive and specific than the other two ratios, with a high AUC. However, for Hirayama disease, the APB-FDI index cutoff value is 11.6, and it shows broad clinical utility with standardized net benefits exceeding those of healthy controls and Hirayama patients when the risk thresholds are between 0.1 and 0.8. The IDI results indicate that the APB-FDI index is superior to the ADM-APB CMAP amplitude ratio and FDI-ADM CMAP amplitude ratio in healthy controls and Hirayama patients.

This study was done among a small number of Bangladeshi ALS patients. NCS parameters may differ in demography and physical factors [20]; we cannot generalize the findings to different races, ages, and physical traits. It needs a multicentered and multinational prospective study to generalize.

In the present study, most ALS patients were 40 years or more, were generally male, and one-third had low body mass index. It is concordant with the demography and characteristics of ALS patients [2,4,21]. Most of the patients were a cervical onset phenotype. The predominant phenotype of ALS is bulbar in some studies [22], but in China dominant type is limb onset [23] in the case of sporadic ALS. As in this study, we studied hand cervical phenotype is frequent here. The patients with Hirayama were generally young and male, as found in the other studies [25,26].

The diagnosis of ALS is not always straightforward. There are no specific biomarkers of ALS, and it is mainly diagnosed by criteria [27,28]. Accurate and early diagnosis is essential to increasing the life expectancy of ALS patients [29]. The most used diagnostic criteria are the revised El Escorial criteria (rEEC) and Awaji criteria (AC), has sensitivity ranges from 40-70%, which increases with the progression of the disease [30]. Electrophysiology may help diagnose early ALS, as some electrophysiologic findings may appear earlier. Spilt hand phenomenon is commonly found across all stages and degrees of certainty of ALS [17,18].

The split hand phenomenon was first described by Wilbourn [10]. Satoshi Kuwabara [18], in their research electro physiologically confirmed the preferential involvement of APB and FDI among ALS patients. In their study, they elucidate APB-ADM, FDI-ADM, and FDI-APB ratio. Among the healthy control, the ratio of APB-ADM, FDI-ADM, and FDI-APB were 1(.3), 1.5(0.4), and 1.6(0.5), respectively [10]. Among the healthy subjects, the APB-ADM ratio was 0.8(0.7-1.2), FDI-ADM was 1.2(0.9-1.4) [18], and both were almost similar. In our study The cutoff values, ADM-APB CMAP amplitude ratio, and FDI-ADM CMAP amplitude ratio are 4.27, 1.04, and 2.20, respectively. In this study, among the ALS patients, the APB-ADM value was 0.6(0.3-1.1), and the FDI-ADM value was 0.8(0.4-1.1). The slight difference in the value might be due to different reporting parameters. Moreover, they used the mean, whereas we used the median. And in our study, the parameter was not normally distributed. They examined the ratio in lower motor neuron disorders (LMND), cervical spondylotic amyotrophy (CSA), and polyneuropathy. They found that the findings are specific to ALS [18].

We found few studies comparing the findings in Hirayama disease, which also presents with the small muscle of hand wasting [31]. This study also found that the split hand phenomenon is unique to ALS. The ratio of APB-ADM and FDI-ADM is reversed in Hirayama disease 6.3(3.2-13,9) and 2(1.2-2.7), respectively. In the case of Hirayama disease, the reverse split hand phenomenon is described in some studies. One study reported APB-ADM ratio 0.89 ± 0.98 [17]. APB amplitude of Hirayama in our study was 6.4(5.3-8.1), similar to Kalita et al. [32]. They found CMAP ratio of ADM-APB was 0.43 (0.29), and in our study, the median value was 0.2(0.1-0.3). Moreover, we found wasting is more prominent in APB than in the FDI (.28 vs. 2.3).

In a study, an ADM-APB ratio >1.7 was found to be 51% sensitive and 99% sensitive [16]; in our study, we found the median ratio of 1.7of (0.9-3.2), and a value >2.2 can diagnose ALS with 21% sensitivity and 96% specificity.

The split hand index was first examined by Menon et al. [11]; they found the index in ALS was 3.5 ± 0.6, SI cutoff value of 5.2 exhibiting a sensitivity of 74% and specificity of 80%. Our study found that the median index was 1.7(0.3-3.8), and a cutoff value lower than 4.2 can diagnose ALS with 72% sensitivity and 100% specificity. In another study, the spilt hand index in Hirayama was found as 13.68 (9.39), whereas in our study, it was 13.6(6.9-15.7), and in ALS, they found it 2.13 (1.90) [32]. In this study, we affirmed the findings of other studies.

We evaluate which ratio performs better using indicators such as AUCs, IDIs, and DCAs. Based on DCA curve analysis, when the threshold probabilities were between 0.5 to 0.7, the order of performance was as follows: APB-FDI, FDI-ADM, and ADM-APB ratios. The APB-FDI index outperformed in all threshold probabilities, but FDI-ADM performed better than ADM-APB after threshold probabilities of 0.5.

Using decision curve analysis (DCA) has several advantages, including the consideration of patient and physician preferences and the use of a threshold probability metric. DCA helps in evaluating the benefits and harms associated with prediction models by assessing the value of correctly treating a positive case versus the risk of treating a false positive case. This makes it an essential tool in clinical decision-making. When determining the risk threshold, a model with a higher standardized net benefit (sNB) is deemed superior [32].

This study found that the APB-FDI index is more effective than other groups, with an AUC of 0.93 and a 95% confidence interval of 0.99-0.98. However, the AUC for FDI-ADM and ADM-APB were only 0.75 and 0.47, respectively. The Youden Index, which summarizes the receiver operating characteristic curve and places equal importance on sensitivity and specificity [33]., showed a high score of 0.72 for the APB-FDI index and a very low score of 0.18 for ADM-APB.

This study found that the overall IDA was less than 0, meaning the model’s performance could have been better. Nevertheless, the APB-FDI index performed slightly better than the other two ratios, although the difference was not significant.

Our research has shown that the APB-FDI index is a more effective diagnostic tool than others, as it simultaneously considers all three muscles’ involvement. Nevertheless, the APB-FDI test may be helpful in cases of severe muscle atrophy and concomitant entrapment neuropathy. It should still be routinely administered to evaluate small muscle wasting in the hands. This can help reduce unnecessary interventions for ALS patients, as research has shown that 5% of ALS patients undergo spinal surgery and 42% have lower back operations[35].

In this study, we provided a comprehensive analysis of the diagnostic performances. However, it is essential to note that our study had certain limitations. The small sample size and the cross-sectional design are inherent limitations that may affect the generalizability of our findings. Additionally, as our data was obtained in real-time, the results may change throughout the disease. Future studies should establish whether this phenomenon is maintained in advanced disease stages through prospective analysis. Unfortunately, due to the limited sample size, we could not conduct subgroup analysis. We recommend further studies comparing this phenomenon in different phenotypes of ALS while also considering the duration of the disease. It should be noted that most patients were evaluated during the routine diagnostic process.

## Conclusion

APB-FDI Index, depicting the split hand phenomenon, is a highly sensitive and specific diagnostic marker and may be helpful in early and conflicting instances to diagnose ALS. Its diagnostic value is also higher than other diagnostic tests, like APB-ADM and FDI-ADM ratio, which reproduce the split hand phenomenon.

## Data Availability

All relevant data are within the manuscript and its Supporting Information files.

## Acknowledgements

We are grateful to every patient who gave their valuable consent for participation in this study; without their help, it would be impossible to conduct this study.

## Acknowledgement

Patients who participate.

## Conflict of Interest

The authors declare that there is no conflict of interests regarding the publication of this paper Research number: ERC-SSMC/2019/224.

## Funding

Received no funds from any source.

## Supporting Information

S1-Correlation between the disease severity of ALS assessed with ALS functional rating scale and the APB-FDI index

## Notes

### Competing Interest Statement

The authors have declared no competing interest.

### Funding Statement

The author(s) received no specific funding for this work.

### Author Declarations

We obtained Ethical clearance from the ethical review committee of Sir Salimullah Medical College Mitford Hospital (SSMC/2019/224).

## References

1. van den Bos, M.A., Geevasinga, N., Higashihara, M., Menon, P. and Vucic, S., 2019. Pathophysiology and diagnosis of ALS: insights from advances in neurophysiological techniques. International Journal of Molecular Sciences, 20(11), p.2818.

2. Coupe, C. and Gordon, P.H., 2013. Amyotrophic lateral sclerosis-clinical features, pathophysiology and management. European Neurol Rev, 8, pp.38–44.

3. Rossi, F. H., Franco, M. C., Estevez, A. G. . Pathophysiology of Amyotrophic Lateral Sclerosis. In: Estévez, A. G., editor. Current Advances in Amyotrophic Lateral Sclerosis [Internet]. London: IntechOpen; 2013 [cited 2022 May 02]. Available from: https://www.intechopen.com/chapters/45326 doi: 10.5772/56562

4. Gordon PH. Amyotrophic Lateral Sclerosis: An update for 2013 Clinical Features, Pathophysiology, Management and Therapeutic Trials. Aging Dis. 2013 Oct 1;4(5):295–310. doi: 10.14336/AD.2013.0400295. PMID: 24124634; PMCID: PMC3794725.

5. Brooks BR. El Escorial World Federation of Neurology criteria for the diagnosis of amyotrophic lateral sclerosis. Subcommittee on Motor Neuron Diseases/Amyotrophic Lateral Sclerosis of the World Federation of Neurology Research Group on Neuromuscular Diseases and the El Escorial “Clinical limits of amyotrophic lateral sclerosis” workshop contributors. J Neurol Sci. 1994 Jul;124 Suppl:96–107. doi: 10.1016/0022-510x(94)90191-0. PMID: 7807156.

6. Brooks BR, Miller RG, Swash M, Munsat TL; World Federation of Neurology Research Group on Motor Neuron Diseases. El Escorial revisited: revised criteria for the diagnosis of amyotrophic lateral sclerosis. Amyotroph Lateral Scler Other Motor Neuron Disord. 2000 Dec;1(5):293–9. doi: 10.1080/146608200300079536. PMID: 11464847.

7. Agosta F, Al-Chalabi A, Filippi M, Hardiman O, Kaji R, Meininger V, Nakano I, Shaw P, Shefner J, van den Berg LH, Ludolph A; WFN Research Group on ALS/MND. The El Escorial criteria: strengths and weaknesses. Amyotroph Lateral Scler Frontotemporal Degener. 2015 Mar;16(1-2):1–7. doi: 10.3109/21678421.2014.964258. Epub 2014 Dec 8. PMID: 25482030.

8. Ghasemi M. Amyotrophic lateral sclerosis mimic syndromes. Iran J Neurol. 2016 Apr 3;15(2):85–91. PMID: 27326363; PMCID: PMC4912674.

9. Paganoni S, Macklin EA, Lee A, Murphy A, Chang J, Zipf A, Cudkowicz M, Atassi N. Diagnostic timelines and delays in diagnosing amyotrophic lateral sclerosis (ALS). Amyotroph Lateral Scler Frontotemporal Degener. 2014 Sep;15(5-6):453–6. doi: 10.3109/21678421.2014.903974. Epub 2014 Jul 1. PMID: 24981792; PMCID: PMC4433003.

10. Wilbourn AJ. The “split hand syndrome”. Muscle Nerve. 2000 Jan;23(1):138. doi: 10.1002/(sici)1097-4598(200001)23:1<138:aid-mus22>3.0.co;2-7. PMID: 10590421.

11. Menon P, Kiernan MC, Yiannikas C, Stroud J, Vucic S. Split-hand index for the diagnosis of amyotrophic lateral sclerosis. Clin Neurophysiol. 2013 Feb;124(2):410–6. doi: 10.1016/j.clinph.2012.07.025. Epub 2012 Sep 25. PMID: 23017503.

12. Eisen A, Kuwabara S. The split hand syndrome in amyotrophic lateral sclerosis. J Neurol Neurosurg Psychiatry. 2012 Apr;83(4):399–403. doi: 10.1136/jnnp-2011-301456. Epub 2011 Nov 19. PMID: 22100761.

13. Kuwabara S, Kanai K. [Altered axonal ion channel function in amyotrophic lateral sclerosis]. Brain Nerve. 2007 Oct;59(10):1109–15. Japanese. PMID: 17969351.

14. Weber M, Eisen A, Stewart H, Hirota N. The split hand in ALS has a cortical basis. J Neurol Sci. 2000 Nov 1;180(1-2):66–70. doi: 10.1016/s0022-510x(00)00430-5. PMID: 11090867.

15. Benny R, Shetty K. The split hand sign. Ann Indian Acad Neurol. 2012 Jul;15(3):175–6. doi: 10.4103/0972-2327.99700. PMID: 22919187; PMCID: PMC3424792.

16. Corcia P, Bede P, Pradat P, et al. Split-hand and split-limb phenomena in amyotrophic lateral sclerosis: pathophysiology, electrophysiology and clinical manifestations. Journal of Neurology, Neurosurgery & Psychiatry 2021;92:1126–1130.

17. GALNARES-OLALDE, Javier A. López-Hernández, Juan C. Saráchaga-Adib, Jorge de. et al. Split hand phenomenon: An early marker for amyotrophic lateral sclerosis. Rev. mex. neurocienc. [online]. 2021, vol.22, n.4, pp.141–145. Epub 30-Jul-2021. ISSN 2604-6180. https://doi.org/10.24875/rmn.20000135

18. Tashiro K, Kikuchi S, Itoyama Y et al., “Nationwide survey of juvenile muscular atrophy of distal upper extremity (Hirayama disease) in Japan,” Amyotrophic Lateral Sclerosis, vol. 7, no. 1, pp.38–45, 2006.

19. Kuwabara S, Sonoo M, Komori T, Shimizu T, Hirashima F, Inaba A, Misawa S, Hatanaka Y; Tokyo Metropolitan Neuromuscular Electrodiagnosis Study Group. Dissociated small hand muscle atrophy in amyotrophic lateral sclerosis: frequency, extent, and specificity. Muscle Nerve. 2008 Apr;37(4):426–30. doi: 10.1002/mus.20949. PMID: 18236469.

20. Cedarbaum JM, Stambler N, Malta E, Fuller C, Hilt D, Thurmond B, Nakanishi A. The ALSFRS-R: a revised ALS functional rating scale that incorporates assessments of respiratory function. BDNF ALS Study Group (Phase III). J Neurol Sci. 1999 Oct 31;169(1-2):13–21. doi: 10.1016/s0022-510x(99)00210-5. PMID: 10540002.

21. Fong SY, Goh KJ, Shahrizaila N, Wong KT, Tan CT. Effects of demographic and physical factors on nerve conduction study values of healthy subjects in a multi-ethnic Asian population. Muscle Nerve. 2016 Aug;54(2):244–8. doi: 10.1002/mus.25029. Epub 2016 May 19. PMID: 26790132.

22. Mariosa D, Beard JD, Umbach DM, Bellocco R, Keller J, Peters TL, Allen KD, Ye W, Sandler DP, Schmidt S, Fang F, Kamel F. Body Mass Index and Amyotrophic Lateral Sclerosis: A Study of US Military Veterans. Am J Epidemiol. 2017 Mar 1;185(5):362–371. doi: 10.1093/aje/kww140. PMID: 28158443; PMCID: PMC5860019.

23. Chen L, Zhang B, Chen R, Tang L, Liu R, Yang Y, Yang Y, Liu X, Ye S, Zhan S, Fan D. Natural history and clinical features of sporadic amyotrophic lateral sclerosis in China. J Neurol Neurosurg Psychiatry. 2015 Oct;86(10):1075–81. doi: 10.1136/jnnp-2015-310471. Epub 2015 Jun 29. PMID: 26124198.

24. Talman P, Duong T, Vucic S, et al. Identification and outcomes of clinical phenotypes in amyotrophic lateral sclerosis/motor neuron disease: Australian National Motor Neuron Disease observational cohort BMJ Open 2016;6:e012054. doi: 10.1136/bmjopen-2016-012054

25. Hirayama K. [Juvenile non-progressive muscular atrophy localized in the hand and forearm--observations in 38 cases]. Rinsho Shinkeigaku. 1972 Jul;12(7):313–24. Japanese. PMID: 4674765.

26. Zhou B, Chen L, Fan D, Zhou D. Clinical features of Hirayama disease in mainland China. Amyotroph Lateral Scler. 2010;11(1-2):133–9. doi: 10.3109/17482960902912407. PMID: 19412815.

27. de Carvalho M, Dengler R, Eisen A, England JD, Kaji R, Kimura J, Mills K, Mitsumoto H, Nodera H, Shefner J, Swash M. Electrodiagnostic criteria for diagnosis of ALS. Clin Neurophysiol. 2008 Mar;119(3):497–503. doi: 10.1016/j.clinph.2007.09.143. Epub 2007 Dec 27. PMID: 18164242.

28. Li TM, Swash M, Alberman E, Day SJ. Diagnosis of motor neuron disease by neurologists: a study in three countries. J Neurol Neurosurg Psychiatry. 1991 Nov;54(11):980–3. doi: 10.1136/jnnp.54.11.980. PMID: 1800671; PMCID: PMC1014620.

29. Rooney J, Byrne S, Heverin M, Tobin K, Dick A, Donaghy C, et al. A multidisciplinary clinic approach improves survival in ALS: a comparative study of ALS in Ireland and Northern Ireland. Journal of neurology, neurosurgery, and psychiatry. 2015;86(5):496–501. pmid:25550416

30. Bademain Jean Fabrice Ido, Imen Kacem, Mahamadi Ouedraogo, Amina Nasri, Saloua Mrabet, Amina Gargouri, Mouna Ben Djebara, Bawindsongré Jean Kabore, Riadh Gouider, “Sensitivity of Awaji Criteria and Revised El Escorial Criteria in the Diagnosis of Amyotrophic Lateral Sclerosis (ALS) at First Visit in a Tunisian Cohort”, Neurology Research International, vol. 2021, Article ID 8841281, 6 pages, 2021. https://doi.org/10.1155/2021/8841281

31. Singh RJ, Preethish-Kumar V, Polavarapu K, Vengalil S, Prasad C, Nalini A. Reverse split hand syndrome: Dissociated intrinsic hand muscle atrophy pattern in Hirayama disease/brachial monomelic amyotrophy. Amyotroph Lateral Scler Frontotemporal Degener. 2017 Feb;18(1-2):10–16. doi: 10.1080/21678421.2016.1223140. Epub 2016 Aug 30. PMID: 27575868.

32. Kalita J, Kumar S, Misra UK, Neyaz Z. Split hand index and ulnar to median ratio in Hirayama disease and amyotrophic lateral sclerosis. Amyotroph Lateral Scler Frontotemporal Degener. 2017 Nov;18(7-8):598–603. doi: 10.1080/21678421.2017.1336561. Epub 2017 Jun 15. PMID: 28616933

33. Zhang Z, Rousson V, Lee WC, Ferdynus C, Chen M, Qian X, Guo Y; written on behalf of AME Big-Data Clinical Trial Collaborative Group. Decision curve analysis: a technical note. Ann Transl Med. 2018 Aug;6(15):308. doi: 10.21037/atm.2018.07.02. PMID: 30211196; PMCID: PMC6123195.

34. Schisterman EF, Faraggi D, Reiser B, Hu J. Youden Index and the optimal threshold for markers with mass at zero. Stat Med. 2008 Jan 30;27(2):297–315. doi: 10.1002/sim.2993. PMID: 17624866; PMCID: PMC2749250.

35. Yoshor, D., Klugh, A., Appel, S. H. & Haverkamp, L. J. Incidence and characteristics of spinal decompression surgery after the onset of symptoms of amyotrophic lateral sclerosis. Neurosurgery. 57, 984–988 (2005).

